# Control dynamics of the COVID-19 pandemic in China and South Korea

**DOI:** 10.1101/2020.05.01.20087650

**Authors:** João Manoel Losada Moreira

## Abstract

Social isolation measures reduce the population’s infection rate with the COVID-19 virus, but its effectiveness is difficult to quantify. The successful control of the pandemic carried out by China and South Korea is studied using a scheme based on the net relative rate of infection which is very sensitive to sudden changes in the epidemic evolution. The net relative rate of infection for China and South Korea without containment measures, that is, with free proliferation of the virus, lies between 10 and 40 %/day or doubling times of infected persons between 3.5 and 2 days. After measures of containment it dropped and stabilized. South Korea stabilized it around 1 %/day and China around 0.05 %/day with doubling times of 70 days and 1400 days, respectively. A discussion is provided about their processes of control and stabilization of the epidemic process and about the scheme used to study them.

## 1. Introduction

Studies are already found in the literature about the dynamics of the COVID-19 pandemic in its early stages in China and its evolution in that country [1–4]. These studies estimated the basic number of reproduction of the infection showing that it was significantly greater than one and indicated that necessity to use social control measures to reduce its infection growth rate. Other works presented events in China until their final control, highlighted the importance of social isolation measures, and estimated that the average number of daily reproduction of infection in Wuhan fell 34 % after the introduction of travel restrictions on 23^th^ January 2020. At the end of March the authors reported a general situation of the spread of the epidemic in several countries, indicating its transformation into a pandemic [5]. The epicenter of the pandemic migrated to Europe and the United States with several countries acknowledging management problems to handle the growing demand of hospital services to treat the COVID-19 patients.

At the end of March, some studies appeared considering processes to contain the pandemic, such as social isolation actions, identification of infection agents, dealing with incomplete information, and about projections and scenarios for the evolution of the epidemic. The basic approach was adopting variations of the SIR model with agents divided into 3 groups: susceptible to infection, infected (infection agents) and recovered (inert agents) [6].

Monitoring exponential processes are common practice in the industry [8,9]. In this work we study evolution of the pandemic in China and South Korea monitoring the exponential evolution of the number of confirmed cases accumulated over time. We take the time series of this variable as the one that can provide information about the dynamic behavior of the pandemic. To interpret the results, a scheme based on the SIR model [6,10] is developed to take into account actions to contain the advance of the pandemic.

This article begins presenting the data, methods and details of the theoretical basis for interpreting the results. Following, we simulate the successful control of the pandemic carried out by China and South Korea and finally we present the conclusions.

## 2. Data and methods

The COVID-19 epidemic spreads across a region and is detected through tests with different efficiencies. The relationship between the total number of infected individuals in a region at day t_n_, I_n_, and the number of infected individuals counted or detected by the tests applied to the population, C_n_, is

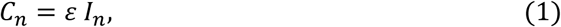

where ε is the overall efficiency of the infection detection system (counts of confirmed cases in the region/true number of infected individuals in the region). If this efficiency is known it is possible to obtain N_n_ over time monitoring C_n_. If the efficiency is unknown, but constant over time, it is possible to obtain true relative rates of change of N_n_ because in the calculation the efficiency cancels out.

Thus, as long as the efficiency ε is invariant over time and adequately covers the region affected by the pandemic (good sample of the universe) the relative infection rate can be estimated correctly through C_n_. If the detection efficiency varies, it is still possible to obtain true net relative rates of infection taking into account such variations. Additionally, ratios such as lethality rate, can provide good estimates of hospital demands and other important time varying information. In this work we take the relative rate of change of C_n_, to model the pandemic temporal evolution.

The data were obtained from the health systems of China and South Korea organized as time series in the Worldometer site [11]. In the rest of the article we use the words pandemic and epidemic interchangeably.

## 3. Temporal model based on the relative rate of change of the number of cases of COVID-19

We consider in the SIR model [6,10] only the balance of the number of infected individuals according to the SIR model), i. e.,

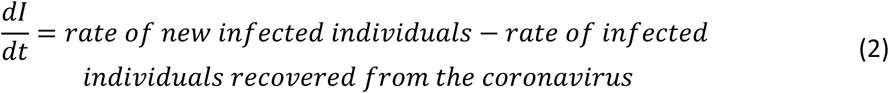

This simplification of the SIR model is due to the fact that the time series for the confirmed number of infected individuals, C_n_, is the most accurate one produced by the coronavirus testing executed the health systems. In this monitoring model, the infection rate by coronavirus is assumed to be proportional to the number of infected individuals on day t, I(t), who act as spreading agents of the infection. We can define a probability of new infection per unit time and per infected individual to determine the infection rate. The rate of removal of infected individuals depends on recovery from the infection, death occurrences or isolation from society. If we assume that this removal rate is also proportional to the number of infected individuals, we can also define a probability of removal of infected individuals per unit time and per infected individual. If we put together the two effects (new infection and removal) we obtain a probability of the net result of infection and removal per unit time and infected individual. Thus Eq. 2 can be written as

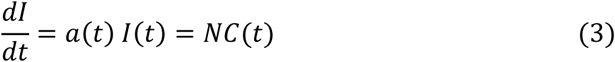

where *a*(*t*) is the probability of net infection accounting for new infection of individuals and removal of infected individuals from society and NC(t) is the net rate of infections appearing in the society (region).

Taking the time interval of one day, the solution of Eq. 3 is given by an exponential evolution of the number of infected individuals, that is,

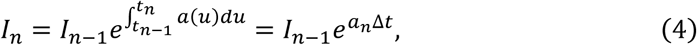

where t_n_ is the n_th_ day, *a_n_* is the average value of a(t) on the n_th_ day, and N_n_ is the number of people infected in the region on day t_n_, and Δt = t_n_-t_n_ −1 or 1 day.

*a_n_* is an indicator of the rate of spread or transmission of the epidemic, because when its value is positive I_n_ grows exponentially, when its value is negative, I_n_ falls exponentially and when it is null I_n_ is constant over time. This probability of net infection per unit of time and per infected individual can be obtained from the total number of confirmed cases time series, C_n_, if the efficiency ɛ of detection of infected individuals is invariant over time and adequately covers the region affected by the epidemic. Thus

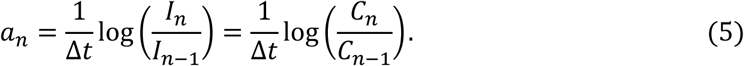

It should be noted that *a_n_* given by Eq. 5 is nothing more than the net relative rate of change in the number of infected individuals calculated considering continuous time variation. In the rest of this article we refer to *a_n_* interchangeably as the net probability of infection per unit of time or net relative rate of infection and is expressed in %/day. The doubling time of infection, td_n_, is related to the net rate of infection as 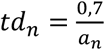.

The main approximation of this model is to consider the efficiency of detection of infections case is approximately constant throughout the study. The number of cases of infection shows great statistical variation at the beginning of the spread of the virus. To reduce the variation, a 3-day moving average was applied to the data before obtaining the net relative infection rates.

## 4. Results and discussion

Figure 1 shows the net relative infection rate for China and South Korea obtained through Eq. 5 using the time series of the number of confirmed cases of infection of these countries [11]. The x-axis shows the days and the day 0 corresponds to 2020/01/31. The y-axis is presented on a logarithmic scale to ease comparing the rates of change in China and South Korea along the studied period. Note that if the efficiency of the detecting systems of both countries has been invariant along the period, Figure 1 shows their true epidemic net relative rate of infection.

**Figure 1 –.**
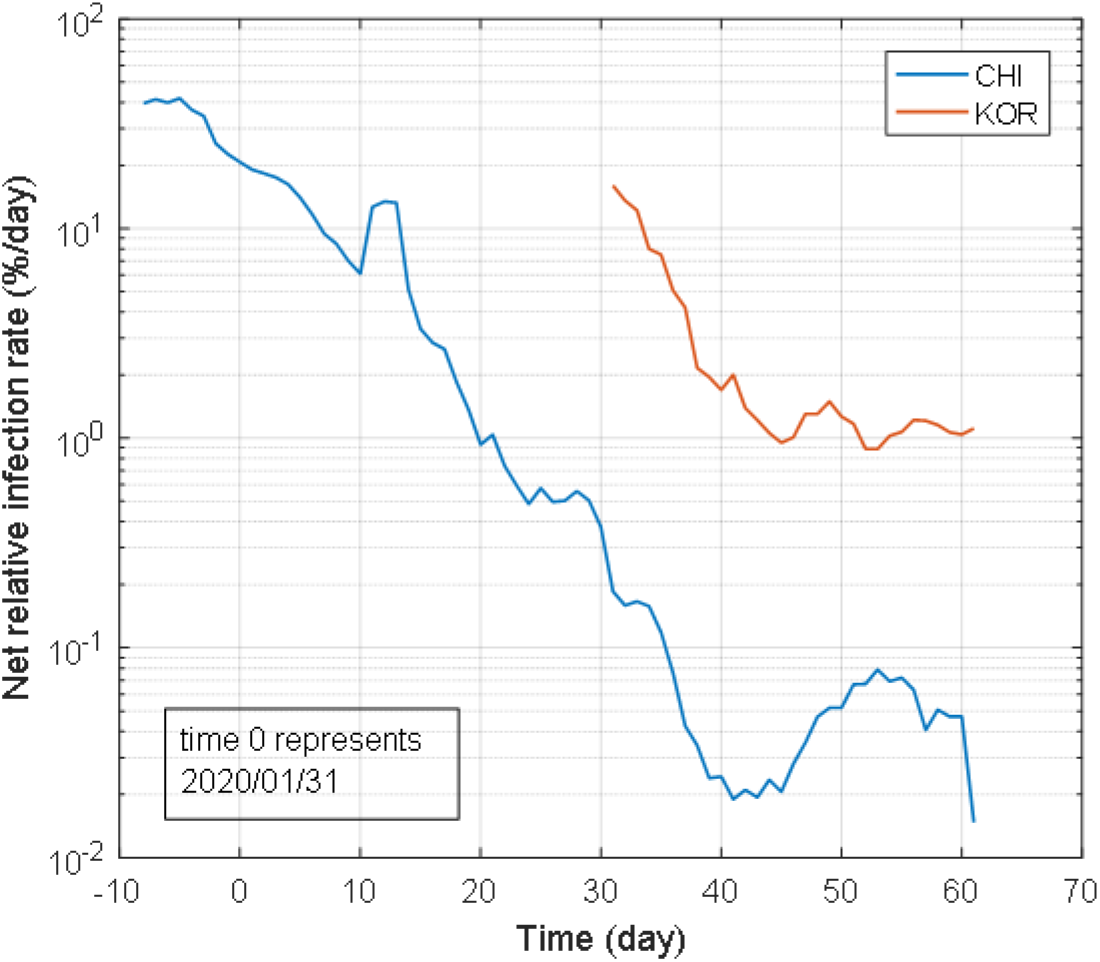
Net relative infection rate as a function of time for China and South Korea.

The net relative rate of infection allows us to monitor the effectiveness of measures to contain the spread of the virus (how fast the relative rate of infection falls). South Korea stabilized around 1 %/day and China around 0.05 %/day. In China, where the epidemic originated, in January 2020 the net probability of infection or net relative rate of infection was 40 %/day and that country started its measures to control the spread of infection in their society. At this time, the doubling time of the number of infected persons was between 2 and 3 days. The measures of social isolation were very strong and produced a continuous reduction of *a_n_*. On February 25^th^, the rate was already less than 1 %/day (70 days of doubling time) and on March 8, less than 0.1 %/day (700 days of doubling time or about 2 years). On 03/27/2020 the confirmed number of infected people in China was 81394, the net relative infection rate was 0.06 %/day and the number of new cases per day, very small.

The infection reached the other countries strongly in March 2020. In South Korea, the infection process started a little earlier and the country took drastic measures to contain the spread of the virus, including social isolation. In early March, South Korea’s net relative infection rate was just below 20 %/day (3.5 days of doubling time). After March 15^th^ it stabilized around 1 %/day (70 days of doubling time) and on 03/27/2020 the confirmed number of infected persons was 9332. Why did this country decide to stabilize its infection rate around 1 %/day and not around 0.05 %/day as China did?

Figure 2 compares the number of new cases per day estimated by Eq. 3 with the new cases obtained from the reported data. NC_n_ given by Eq. 3 is a good approximation of the true number of new confirmed cases per day. The mean relative discrepancy for China was 4.8 % and for South Korea, 12 %.

**Figure 2 -.**
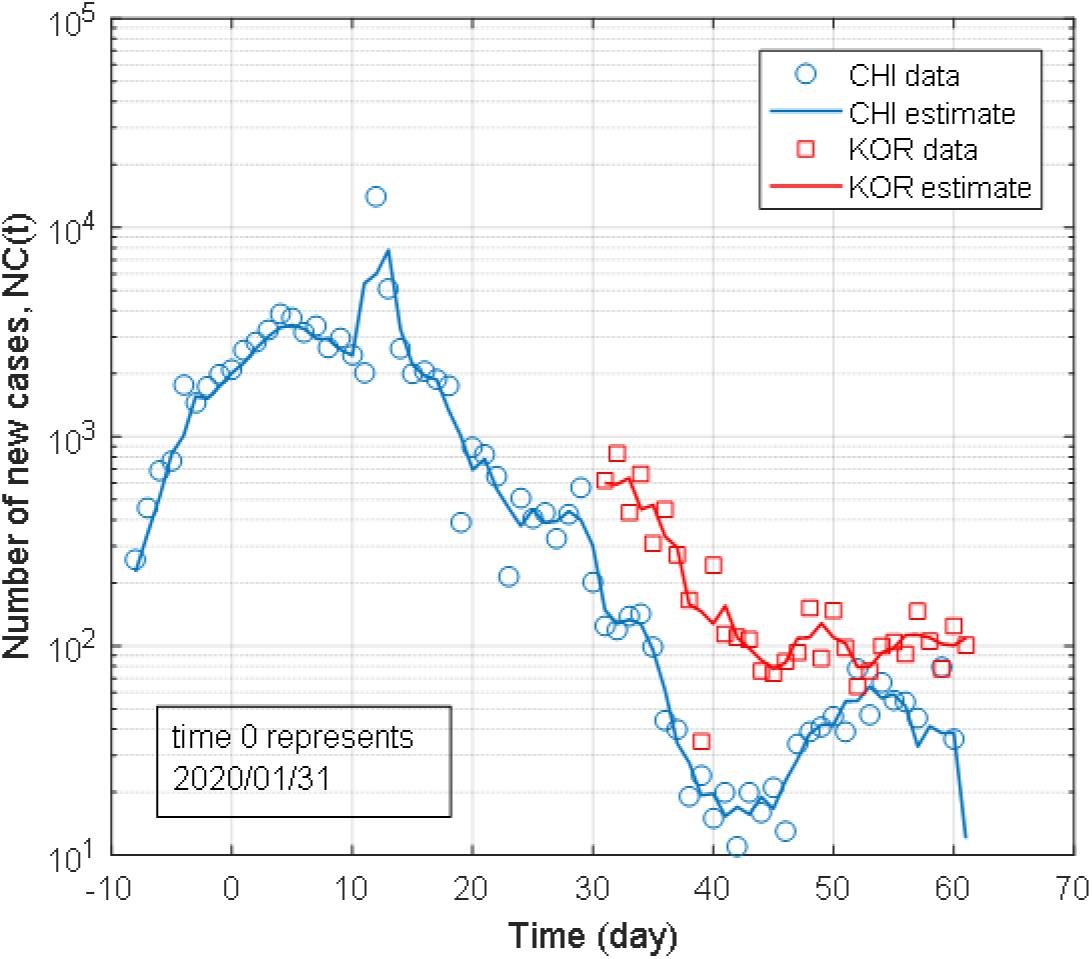
Comparison between the number of detected new cases reported in the data and the rate of detected new cases estimated by Eq. 3.

Figure 2 shows that between 50 and 60 (2^nd^ half of March) the number of new cases in China was around 55 and in South Korea, around 90. These figures were already very similar although the number of confirmed cases in China was 10 times greater. The answer to the above question is that in China the net probability of infection per unit of time was 10 times smaller than that in South Korea. The important variable was NC_n_, the rate of new confirmed cases per day, obtained from the product *a_n_C_n_* which can be directly correlated to hospital demands. A figure between 50 and 100 new confirmed cases per day was suitable for their health and epidemic control systems.

More importantly, Figure 1 shows that the logarithm of the net relative rate of infection is very sensitive to sudden changes of the epidemic evolution. Thus it can be used to monitor the effectiveness of containment actions of the epidemic. Additionally, it presents clear trends of the epidemic process and thus can be used to project future scenarios for the epidemic evolution.

## 5. Conclusions

The evolution of the control process of the pandemic in China and South Korea was presented using a scheme based on the net relative rate of infection. For these two countries, the net rate of infection without containment measures, that is, with free proliferation of the virus, is between 20 and 40 %/day or doubling times of infected persons between 3.5 and 2 days.

After actions to contain the spread of the virus were applied this parameter started to fall indicating that the containment actions taken to control the pandemic were effective. The stabilization limit depends on the capacity of each country to take care of its fellow citizens and the confirmed number of persons infected. If the latter grows significantly, the effort to contain the spread of the virus is greater. South Korea stabilized the process in ~ 15 days and China in ~ 50 days.

The scheme is not a model that simulates the epidemic but a monitoring scheme that infer the current net relative rate of infection based on reported confirmed cases of infection. The logarithm of the net relative rate of infection is very sensitive to sudden changes in the epidemic evolution and can be used to monitor the effectiveness of containment actions of the epidemic. Additionally, it presents clear trends of the epidemic process and can be used to project future scenarios for the epidemic evolution.

## Data Availability

All data utilized in the manuscript are available.

## Acknowledgments

The author thanks the Brazilian agency Coordenação de Aperfeiçoamento de Pessoal de Nível Superior (CAPES) for financial support and acknowledges it had no interference on data collection, interpretation, or decision to submit the work for publication.

## Notes

### Competing Interest Statement

The authors have declared no competing interest.

